# Beta-blocker exposure is associated with nonunion in a geriatric cohort of 253,266 extremity fractures

**DOI:** 10.1101/2023.07.14.23292608

**Authors:** Lillia Steffenson, Brook Martin, Adam Kantor, Dillon O’Neill, Luke Myhre, Tyler Thorne, David Rothberg, Thomas Higgins, Justin Haller, Lucas Marchand

## Abstract

Previously published animal studies have shown positive skeletal effects with local or systemic administration of beta blockers (BBs). However, population studies have shown mixed effects on bone mineral density (BMD) and fracture risk with BB use. The goal of this study was to evaluate whether exposure to BB is associated with fracture nonunion. Fee-for-service Medicare beneficiaries with an extremity fracture were identified by International Classification of Diseases (ICD)-10 and current procedural terminology (CPT) codes from 2016-2019. Charlson Comorbidity Index (CCI) was assigned using diagnoses prior to index fracture and nonunion identified by ICD-10 or CPT codes within one year from index fracture diagnosis. Patients were classified by BB exposure based on Part D (Pharmacy) claims between 90 days prior to and one year following index fracture. Chi square and Student’s T-tests were performed on categorical and continuous variables, respectively. Logistic regression was performed to evaluate the association between BB use and nonunion, controlling for age, sex, race, and comorbidity. Total number of fractures meeting inclusion criteria was 253,266 with 45% of patients having used a BB during the study period. The incidence of nonunion was 3.9% overall. BBs were associated with a 13% increase in non-union for all fracture types, after controlling for age, sex, fracture location, and CCI (OR 1.13 [CI 1.06-1.20], p<.001). Results of this study suggest a negative influence of BB on bone healing, contrary to results of previously published animal models and epidemiologic observations, and demonstrate that BB use during fracture care is associated with significant increase in incidence of nonunion.

## INTRODUCTION

Beta blockers are a class of medications which act as antagonists to beta-adrenergic receptors of the sympathetic nervous system (SNS). These receptors are found primarily in mesenchymal tissue types including cardiac, pulmonary, and vascular smooth muscle as well as skeletal muscle, adipose and osseous tissue. There are three subtypes of this adrenergic stimulated G-protein coupled receptor, beta-1, beta-2, and beta-3 receptor, and effects vary due to the distribution of these receptors in different tissues. Beta-1 receptors found within the heart increase heart rate and contractility causing a rise in cardiac output while those in the kidneys promote renin secretion. When stimulated, beta-2 receptors result in bronchodilation, vasodilation, and enhanced skeletal muscle contraction in addition to gluconeogenesis and insulin secretion. Beta-3 receptors within fat tissue increases lipolysis and relaxation of bladder smooth muscle.

Skeletal homeostasis is maintained by the constant balance of activity between osteoblasts (bone forming cells) and osteoclast (bone resorbing cells). This mechanism may alter bone mineral density due to changes in activity or underlying physiologic demands (1, 2). Within osseous tissue, beta-2 adrenergic receptors (β2AR s) are found on both osteoblasts and osteoclasts where β2AR activation by norepinephrine and epinephrine induces bone loss through RANK ligand-mediated activation of osteoclasts and inhibits osteoblast proliferation (3). In mice, deletion of adrenergic beta-2 receptor (Adrb2) leads to greater trabecular bone microarchitecture and bone mass within both femur and vertebrae. Addition of beta-1 receptor knockout to the model results in loss of this phenotype and overall decreased total bone volume and cortical thickness (4) suggesting differential effects of these two receptor subtypes.

Observational studies in humans have similarly identified that individuals taking beta blocker medications have higher bone mineral density (BMD) and lower fracture risk (5–13). Considering post-menopausal women have been found to have higher sympathetic activity compared to pre-menopausal women and sympathetic nerve activity is inversely correlated with trabecular bone volume (14), these findings have renewed interest in modulating adrenergic signaling as a potential intervention for primary osteoporosis prevention (2). One such study by Khosla et al., saw increases in distal radius BMD in post-menopausal women treated with 20 weeks of atenolol (15).

At present, there are no human studies evaluating the effect of beta blockers on fracture outcome. However, in a murine fracture model, low dose propranolol over 9 weeks increased periosteal and endosteal bone formation and improved consolidation of fracture callus. In the absence of fracture, torsional strength of rat femurs was increased by 33% with propranolol (16). In another study, intraperitoneal administration of the cardio selective BB nebivolol, had a dose dependent increase in histologic osseous healing in rat femur fractures stabilized with intramedullary Kirschner wires, whereas cartilage formation was greater in the control group (17). However, rat osteotomy healing with intraperitoneal propranolol administration in a third study did not change final torsional strength or union rates, despite a nonsignificant increase in callus strength at 5 weeks in the propranolol group (18).

The purpose of the present study was to evaluate the association between beta blocker exposure and fracture healing in humans, given the evidence for anabolic osseous effect in prior observational and animal studies.

## METHODS

### Study Cohort

A retrospective analysis of a convenience cohort drawn from Medicare beneficiaries with a Part B (provider service) claim for back pain and/or osteoporosis from 2016-2019 was performed. Patients were included if they were over 65 years of age and had a Part B inpatient or outpatient claim for a femur, tibia/fibula, forearm, or humerus fracture identified by International Classification of Diseases and 10^th^ Revision (ICD-10) codes and Current Procedural Terminology codes (CPT) (Supplemental Table 1). From this initial cohort, patients were then excluded if they were less than 65 years old at the time of fracture, had Medicare through either the end stage renal disease (ESRD) or Social Security Disability Insurance (SSDI) entitlement programs, were not enrolled in Medicare Part D coverage, had follow-up of less than 1 year (e.g. fracture occurred in 2019), died within a year of fracture, had a concurrent or prior diagnosis of nonunion by ICD-10 codes at the time of index fracture, or had insufficient 1 year claims history prior to fracture to calculate comorbidity index. The cohort was screened for duplicate claims which were excluded from final analysis. Patient demographics including age, sex, race (simplified grouping as “White”, “Black”, or “Other”), and Charlson Comorbidity Index (CCI) (19) were collected.

The primary outcome of this study was fracture status one year following index injury, dichotomized as union or non-union. Non-union was identified in the final cohort using CPT codes for nonunion and ICD-10 codes (Supplemental Table 1). A union of the fracture was assumed if the beneficiary claims history did not include a non-union diagnosis.

The analytical cohort was grouped based on exposure to beta blockers (supplemental table 2) and statin medications for comparison and categorized as pre-fracture use only, post-fracture use only, or both pre- and post-fracture exposure. Pre-fracture exposure was defined as a prescription claim within 90 days prior to index fracture. Subgroup analyses were also performed to evaluate differences in immediate post-fracture exposure within 90 days of index fracture, difference in non-union within each specific type of fracture, between selective vs. non-selective beta blockers, then beta blockers were combined with statins, and based number of beta blocker (statin) prescriptions filled served as a proxy for an exposure “dose-response”,

### Statistical Analysis

Descriptive statistics were presented for patient demographics, fracture types, rates of union and non-union, and medication use. Chi square and Student’s T-tests were performed on categorical and continuous variables, respectively. Separate multivariable logistic regression models with robust standard errors were then used to describe the association between each medication class and fracture status at 1-year, both overall and within each fracture type, controlling for age, sex, race and CCI. Similar logistic regression models were used to examine the relationship between medication timing and non-union while controlling for age, sex, race, CCI, and fracture type. Analyses were conducted using Stata 17.0 MP (College Station, TX) accessed through Medicare’s Virtual Research Data Center, and presented as odds ratios with hypotheses based on an alpha level of 0.001. A power analysis performed using a t beta value of 0.80 indicated at least 5300 patients needed in each group to detect a 1% difference in nonunion.

## RESULTS

Starting with 1,079,157 extremity fractures, a total of 253,266 fractures met inclusion criteria (Figure 1) with 85,114 femur fractures, 73,444 forearm fractures, 41,097 humerus fractures and 53,373 tibia or fibula fracture. Overall incidence of nonunion was 3.9%. The highest incidence of nonunion was observed among humerus fractures at 6.2%. Examining patient demographics, those diagnosed with nonunion were younger by an average of 2 years (p<.001) and there was a higher proportion of males (21.3% vs 17.8%, p<.001) (Table 2). There were no significant differences in race or CCI by fracture outcome.

**Figure 1.**
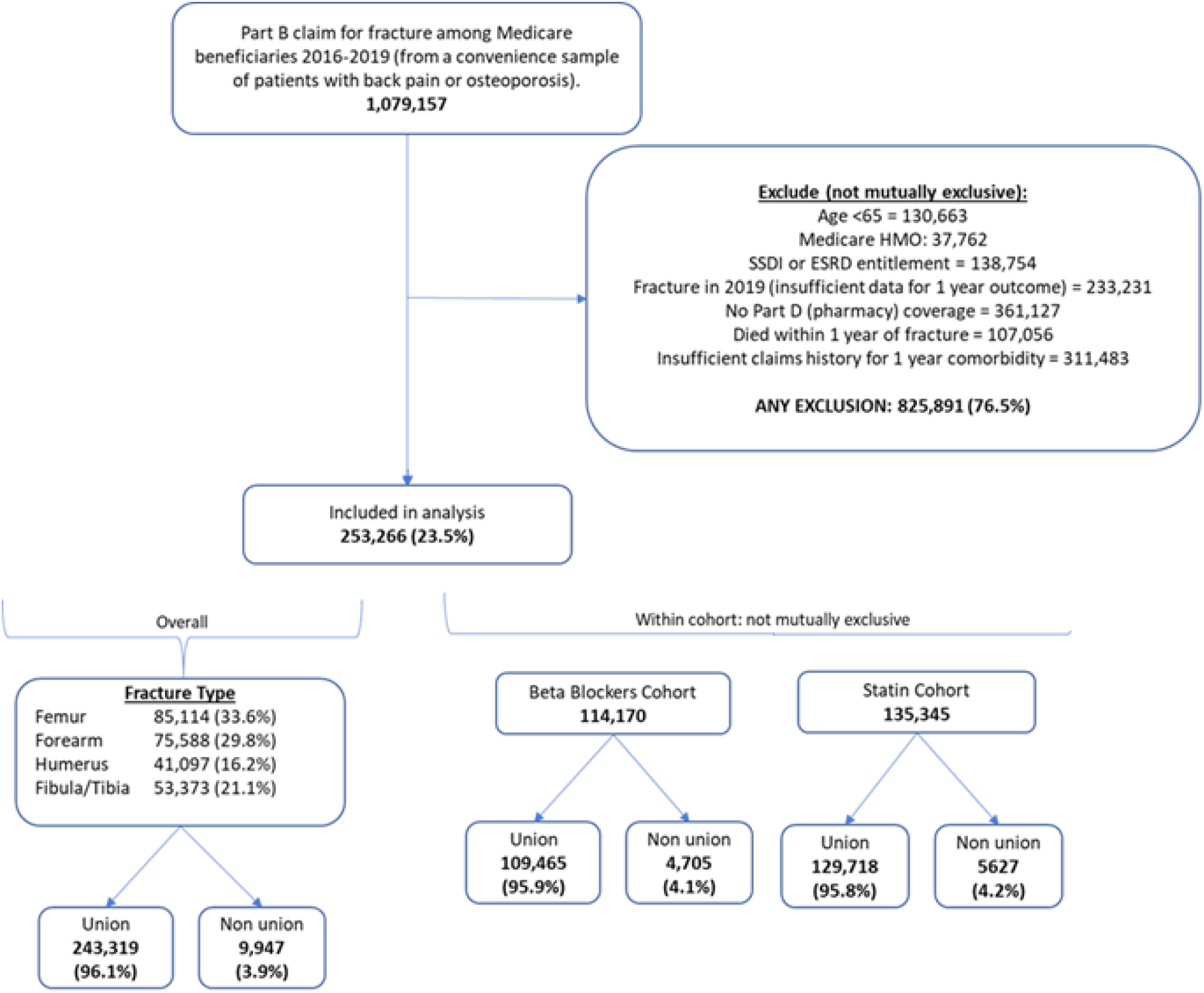
Consort diagram of study cohort. Patients were excluded if they were eligible for Medicare through social security disability (SSDI), end-stage renal disease (ESRD) entitlements, had insufficient Part D pharmacy data or at least 1 year of claims data.

**Table 1.** Fracture union status at 1-year, within fracture type cohort.

| Fracture Cohort | Union | Non union | Total |
| --- | --- | --- | --- |
| Femur | 82,059 (96.4%) | 3,055 (3.6%) | 85,114 |
| Forearm | 73,444 (97.2%) | 2,144 (2.8%) | 75,588 |
| Humerus | 38,566 (93.8%) | 2,531 (6.2%) | 41,097 |
| Fibula/Tibia | 51,041 (95.6%) | 2,332 (4.4%) | 53,373 |

**Table 2.** Demographic Characteristics between Medicare beneficiaries who did or did not experience non-union within one year following fracture.

|  | Fracture outcome at 1 year |  |  | p-value |
| --- | --- | --- | --- | --- |
|  | Union<br>(n=243,319) | Non-union<br>(n=9,947) | Total<br>(n=253,266) |  |
| Age (mean) | 79.4 | 77.3 | 79.3 | <0.001 |
| Female, % | 82.2 | 77.7 | 82.0 | <0.001 |
| Race (3 groups), % |  |  |  |  |
| White | 91.6 | 91.6 | 91.6 | 0.407 |
| Black | 3.3 | 3.1 | 3.2 |  |
| Other | 5.2 | 5.4 | 5.2 |  |
| Comorbidity 365-day lookback, % |  |  |  |  |
| 0 | 21.7 | 21.4 | 21.7 | 0.739 |
| 1 | 20.6 | 20.5 | 20.6 |  |
| 2 | 57.8 | 58.1 | 57.8 |  |
| Medication use |  |  |  |  |
| Beta blocker, % | 45.0 | 47.3 | 45.1 | <0.001 |
| Statin, % | 53.3 | 56.6 | 53.4 | <0.001 |

Exposure to beta blockers and statins was greater in males, patients with higher CCI, and black race (Table 3).

**Table 3.** Demographic Characteristics between Medicare beneficiaries based on medication exposure.

|  |  | **Beta Blocker** |  | **p value** | **Statin** |  | **p value** |
| --- | --- | --- | --- | --- | --- | --- | --- |
|  |  | No | Yes |  | No | Yes |  |
| **N** |  | 139,096 | 114,170 |  | 117,921 | 135,345 |  |
| **Age, mean (sd)** |  | 78.6<br>(8.1) | 80.2<br>(7.8) |  | 79.9<br>(8.5) | 78.9<br>(7.6) |  |
| **Female, %** |  | 83.1 | 80.7 | p<.001 | 84.8 | 79.6 | p<.001 |
| **Race (3 groups), %** |  |  |  |  |  |  |  |
|  | White | 91.6 | 91.6 |  | 92.2 | 91 |  |
|  | Black | 2.9 | 3.6 |  | 3 | 3.4 |  |
|  | Other | 5.5 | 4.8 | p<.001 | 4.8 | 5.5 | p<.001 |
| **Comorbidity 365-day lookback, %** |  |  |  |  |  |  |  |
|  | 0 | 27.8 | 14.2 |  | 27.8 | 16.3 |  |
|  | 1 | 22.9 | 17.6 |  | 22.5 | 18.9 |  |
|  | 2 | 49.2 | 68.2 | p<.001 | 49.7 | 64.8 | p<.001 |

After controlling for age, sex, race, comorbidity, and fracture type, the multivariable model did not reveal a significant association between statin use and nonunion (OR 1.05 [95%CI 1.01 – 1.47] p-value=0.018). However, beta blockers were associated with an 11% increase in nonunion diagnosis (OR 1.11 [95%CI 1.06-1.15] <0.001) regardless of the number of prescriptions in the follow up period (Table 4). When excluding those taking statins, the increase of nonunion among beta blocker users was 13% (OR 1.13 [95% CI 1.06-1.20]).

**Table 4.** Association of select medications on non-union, controlling for age, sex, race, Charlson comorbidity index (365-day lookback), and fracture type. (Prescription = Rx)

| Model | Factor | OR (95%CI) | Rate | p-value |
| --- | --- | --- | --- | --- |
| Non-union as a function of any exposure to medication within 1 year of fracture. | Beta-blocker | 1.11 (1.06 – 1.15) | 3.7 vs.3.4 | <0.001 |
|  | Statin | 1.05 (1.01 – 1.10) | 3.6 vs 3.5 | 0.018 |
| Non-union as a function of beta blocker prescriptions filled (dose response) conditional on beta blocker use | 1-4 Rx | 1.00 (ref) | 4 | -- |
|  | 5-6 Rx | 0.96 (0.89 – 1.03) | 3.8 | 0.258 |
|  | 7+ Rx | 0.93 (0.87 – 1.00) | 3.7 | 0.045 |
| Non-union as a function of statin prescriptions filled (dose response) conditional on statin use | 1 -4 Rx | 1.00 (ref) | 4.1 | -- |
|  | 5 Rx | 0.88 (0.83 – 0.95) | 3.6 | 0.001 |
| Non-union as a function pre and post-fracture beta blocker use | None | 1.00 (ref) | 3.5 | -- |
|  | Pre fracture beta-blocker use only | 1.15 (0.97 – 1.35) | 4.0 | 0.110 |
|  | Post fracture beta-blocker use only | 1.14 (1.07 – 1.22) | 3.9 | <0.001 |
|  | Pre and post fracture beta-blocker use | 1.12 (1.07 – 1.17) | 3.9 | <0.001 |

### Subgroup analysis

There was no obvious pattern in the likelihood of non-union based on the timing of beta-blocker use relative to when the fracture occurred (Table 4). There was no difference between selective (OR 1.12 [95%CI 1.07 – 1.17]), non-selective (OR 1.15 [95%CI 1.07 – 1.23]) or combined selective and non-selective (OR 1.15 [95%CI 0.99 – 1.34] BB exposure on fracture status at 1 year (Table 5). All three categories had a higher risk compared to the non-user group. Combined exposure of beta blocker and statin use had a modest increase in nonunion (OR 1.19 [1.12-1.25 p<0.001] compared to non-users but was not significantly more than taking either beta-blockers or stains alone (Table 5). Neither a greater number of beta blockers prescription nor statin prescriptions filled was associated with a greater likelihood of non-union (Table 4).

**Table 5.** Sub analysis of combined medication exposure and beta blocker class on non-union, controlling for age, sex, race, Charlson comorbidity index (365-day lookback), and fracture type.

| Model | Factor | OR (95%CI) | Rate | p-value |
| --- | --- | --- | --- | --- |
| Non-union as a function of interaction between beta-blocker use and statin use | None | 1.00 (Ref) | 3.4 | -- |
|  | Beta-blocker | 1.13 (1.06 – 1.20) | 3.8 | <0.001 |
|  | Statin | 1.08 (1.02 – 1.14) | 3.6 | 0.009 |
|  | Beta-blocker & statin | 1.19 (1.12 – 1.25) | 3.9 | <0.001 |
| Non-union as a function of beta-blocker type (selective or non-selective), conditioned on use of beta blocker | Select beta-blocker | 1.00 (Ref) | 3.8 | - |
|  | Non-select beta-blocker | 1.03 (0.96 – 1.11) | 3.9 | 0.355 |
|  | Select & non-select beta-blocker | 1.04 (0.89 – 1.21) | 4 | 0.621 |

### Sensitivity analysis for unobserved confounding

We performed a sensitivity analysis to explore the possibility for unobserved confounders to account for the higher risk of non-union found among beta-blocker users at 1 year. By definition, a confounder is a factor associated with both the exposure (beta blocker use) and the outcome (fracture non-union). A confounder sensitivity analysis reports the magnitude that these associations require in order to nullify the association that we observed. In our analysis, we found that a hypothetical unobserved factor with an OR of 1.46 [or a lower CI limit of 1.31] for an association with both the exposure and outcome was required to render our findings for beta blockers null. As shown in Figure 2, a lesser association with the exposure requires an exponentially greater association with the outcome (and vice versa).

**Figure 2.**
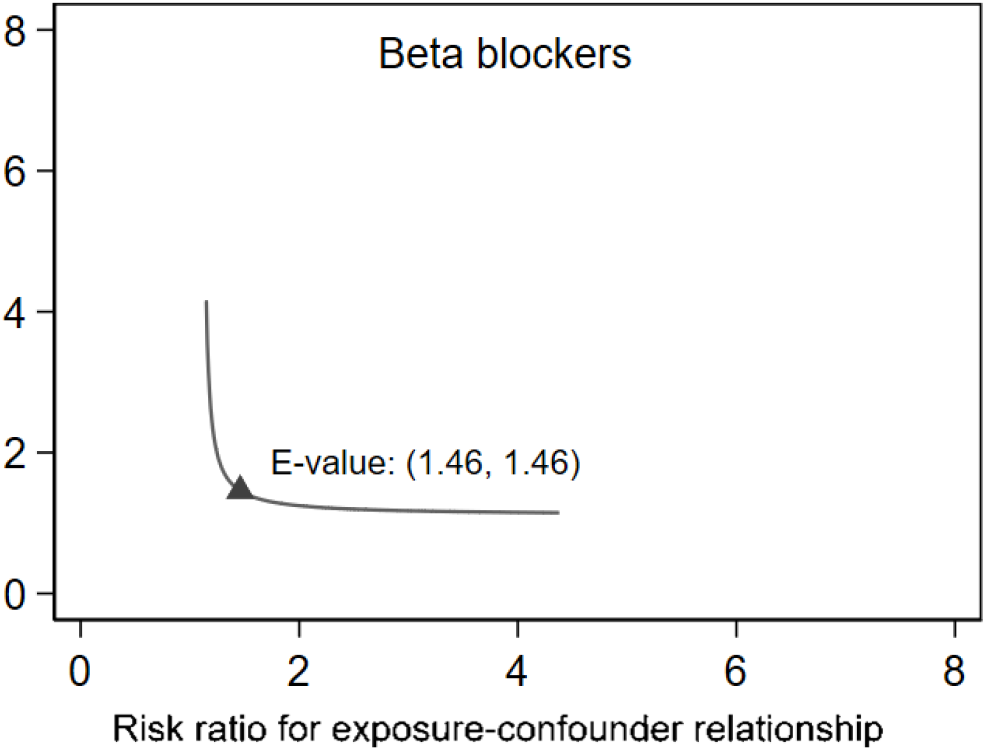
Sensitivity analysis examining the relationship between potential unaccounted confounders.

## DISCUSSION

Age-related bone loss is multi-factorial. The primary mechanism in both men and women is gonadal sex steroid deficiency (20). While this change is more pronounced in post-menopausal women, men also experience age related declines in bone density (21, 22). In women this can be attenuated by hormone replacement therapy (HRT) (23–25), due to HRT stimulation of growth hormone, TGFB and IGF-1 production (26–28). Use of HRT to treat osteopenia or osteoporosis, however, is associated with increased risk of cardiovascular events and breast cancer and is not recommended as first line therapy (21). Geriatric patients also have increased sympathetic nervous system activity, contributing to the higher risk of multi organ system pathophysiologic changes with aging (29, 30) as well as age-related bone loss (14).

Multiple epidemiologic studies from 2000-2015 identified that patients taking BBs have higher BMD and lower fracture risk (5–13). Preceding data from murine models also found an anabolic effect of beta blockers on bone, with osteotomized rats exposed to BB having greater callus formation and improved biomechanics compared to controls (16, 17). The proposed mechanism of this observation is antagonism of normal skeletal B2AR function in stimulating osteoclasts (31). While some animal studies similarly found positive effects of beta blockers on osteotomy healing, there is competing basic science evidence for this association across rodent models (18, 32–35). The aim of this study was to further evaluate the association between beta blockers and fracture fracture healing in humans.

In the present study, there was a modest but significant increase in the diagnosis of nonunion among patients taking beta blockers. There was no difference between cardioselective and nonselective beta blocker exposure. This finding is supported by a previous database study, which also observed an association between cardiac medications and diuretics and nonunion (36). The present study focused on beta blocker use given the abundance of basic science research regarding the mechanism of action of beta blockers in modulating osseous physiology. Notably, the study by Buccheit et al., did not detect higher incidence of nonunion with steroid medications or immunosuppressant exposure following fracture treatment.

Our findings contradict the existing literature regarding the osseoanabolic effect of beta blockers and ongoing research regarding beta blockers as a potential primary preventative agent for osteopenia and osteoporosis (2). However, it is important to consider that adrenergic receptors have differential effects depending on the subtype targeted by pharmacologic interventions. For example, the primary subtype expressed in bone is β2AR, which when stimulated by glucocorticoids results in lower bone density, while co-treatment with propranolol was protective against bone loss (37). On the other hand, beta-1 receptor knockout mice at 4 months have lower BMD, lower bone volume, and trabecular volume when compared to both wild type (WT) and Adbr2 knockout mice (4), indicating that normal adrenergic beta-1 receptor function is important for maintenance of bone volume and density in a murine model. Furthermore, in the same study by Pierroz et al., Adbr1b2^-/-^ mice had persistent decreases in whole body BMD up to 40 and 50 weeks of age, but this difference diminished as the mice approached 60 weeks of age. Based on these findings, the authors propose that beta 2 signaling is higher in bone of wild type mice, but beta-1 signaling may be just as important in the regulation of skeletal tissue.

Additionally, one must consider the condition where SNS activation is known to improve fracture healing. In moderate and severe traumatic brain injury (TBI), fracture healing is accelerated in both clinical studies and murine models (38–41). TBI results in several pathophysiologic effects, but largely increases SNS activity. In an unpublished independent study by Jahn et al., the authors found that increased NE-Arb2 signaling mediates TBI-induced skeletal manifestations in both bone remodeling and regeneration. Furthermore, the study demonstrates that Adrb2 plays an age-dependent and pharmacologically targetable role in fracture healing by promoting callus vascularization under conditions of increased sympathetic tone. The beneficial effect of TBI on bone regeneration was also completely absent in mice lacking Adrb2 (*medRxi*v BIORXIV/2023/548550). These findings suggest an essential function of Adrb2 in promoting bone repair and provide evidence that in a hyperadrenergic state, activity by these receptors may be essential for normal bone healing.

The challenge in these large observational studies remains the difficulty in determining the medication effect, versus a reflection of the patient’s overall health status that is not completely captured by comorbidity indices. The studies in question all use different methods to control for comorbidities, which may confuse the results. It is also difficult to discern if route of medication administration and age of the specimens may change the central and peripheral effects of these medications. Finally, the present study is limited as it is a convenience cohort of Medicare patients is not generalizable to the general population.

## Data Availability

All data produced in the present study are available upon reasonable request to the authors

## AUTHOR CONTRIBUTIONS

Authorship note: LS and BM collaborated on study design, LM supervised the study, BM performed statistical analysis, LS prepared the manuscript which was reviewed and edited by the remaining authors.

## ACKNOWLEDEMENT

Funding support provided by internal grant from the LS Peery MD Discovery Program of the University of Utah’s Department of Orthopaedics.

