## Supplemental table 1 and 2 for "Beta-blocker exposure is associated with nonunion in a geriatric cohort of 253,266 extremity fractures"

**Supplemental table 1: ICD-9/10 and CPT codes for extremity fractures**

| Bone | Procedure Description | CPT Code | ICD 9 Procedure | ICD 10 Procedure | ICD 9 Diagnosis | ICD 10 Diagnosis |
|---|---|---|---|---|---|---|
| Humerus | Fracture | 23600 | 78.12 | 0PH!0*Z | 812.+ | S42.20+ |
| | Operative | 23605 | 78.52 | 0PH!3*Z | | S42.21+ |
| | Fixation | 23615 | 79.00 | 0PH!4*Z | | S42.22+ |
| | | 24500 | 79.01 | 0PS!*4Z | | S42.23+ |
| | | 24505 | 79.11 | | | S42.24+ |
| | | 24515 | 79.21 | ! = C, D, F, G | | S42.29+ |
| | | 24516 | 79.31 | * = 4, 5, 6, B | | S42.30+ |
| | | 24530 | 79.41 | | | S42.32+ |
| | | 24535 | 79.51 | | | S42.33+ |
| | | 24538 | 79.61 | | | S42.34+ |
| | | 24545 | 79.91 | | | S42.35+ |
| | | 24546 | | | | S42.36+ |
| | | 24575 | | | | S42.39+ |
| | | 24576 | | | | S42.40+ |
| | | 24577 | | | | S42.41+ |
| | | 24579 | | | | S42.42+ |
| | | 24582 | | | | S42.45+ |
| | | 24586 | | | | S42.46+ |
| | | | | | | S42.47+ |
| | | | | | | S42.49+ |
| | Nonunion | 24430 | | | 733.82 | S42.20%K |
| | | 24435 | | | | S42.21%K |
| | | | | | | S42.22%K |
| | | | | | | S42.23%K |
| | | | | | | S42.24%K |
| | | | | | | S42.29%K |
| | | | | | | S42.30%K |
| | | | | | | S42.32%K |
| | | | | | | S42.33%K |
| | | | | | | S42.34%K |
| | | | | | | S42.35%K |
| | | | | | | S42.36%K |

| | | | | | | | | |
|---|---|---|---|---|---|---|---|---|
| | | | | | | | S42.39%K<br>S42.40%K<br>S42.41%K<br>S42.42%K<br>S42.45%K<br>S42.46%K<br>S42.47%K<br>S42.49%K<br><br>%= 0-9 | |
| Forearm | Fracture<br>Operative<br>Fixation | 24620<br>24635<br>24650<br>24655<br>24670<br>24675<br>24685<br>25360<br>25500<br>25505<br>25515<br>25520<br>25525<br>25526 | 25530<br>25535<br>25545<br>25560<br>25565<br>25574<br>25575<br>25600<br>25605<br>25606<br>25607<br>25508<br>25609 | 78.13<br>78.53<br>79.02<br>79.12<br>79.22<br>79.32<br>79.42<br>79.52<br>79.62<br>79.92 | 0PH$*4Z<br>0PS$*4Z<br>0PH$*4Z<br>0PS$*4Z<br><br>$ = H, J, K, L<br>* = 0, 3, 4 | 813.+ | S52.00+<br>S52.02+<br>S52.03+<br>S52.04+<br>S52.09+<br>S52.10+<br>S52.12+<br>S52.13+<br>S52.18+<br>S52.20+<br>S52.22+<br>S52.23+<br>S52.24+<br>S52.25+<br>S52.26+<br>S52.27+<br>S52.29+<br>S52.30+<br>S52.32+ | S52.33+<br>S52.34+<br>S52.35+<br>S52.36+<br>S52.37+<br>S52.39+<br>S52.50+<br>S52.53+<br>S52.54+<br>S52.55+<br>S52.56+<br>S52.57+<br>S52.59+<br>S52.60+<br>S52.69+<br>S52.90X+<br>S52.91X+<br>S52.92X+ |

| | | | | | | | |
|---|---|---|---|---|---|---|---|
| | Nonunion | 25400<br>25405<br>25415<br>25420 | | | 733.82 | S52.00%@<br>S52.02%@<br>S52.03%@<br>S52.04%@<br>S52.09%@<br>S52.10%@<br>S52.12%@<br>S52.13%@<br>S52.18%@<br>S52.20%@<br>S52.22%@<br>S52.23%@<br>S52.24%@<br>S52.25%@<br>S52.26%@<br>S52.27%@<br>S52.29%@<br>S52.30%@<br>S52.32%@<br><br>%= 0-9<br>@=K, M, N | S52.33%@<br>S52.34%@<br>S52.35%@<br>S52.36%@<br>S52.37%@<br>S52.39%@<br>S52.50%@<br>S52.53%@<br>S52.54%@<br>S52.55%@<br>S52.56%@<br>S52.57%@<br>S52.59%@<br>S52.60%@<br>S52.69%@<br>S52.90X@<br>S52.91X@<br>S52.92X@<br><br>%= 0-9<br>@=K, M, N |
| Femur | Fracture<br>Operative<br>Fixation | 27235<br>27238<br>27240<br>27244<br>27245<br>27246<br>27500<br>27501<br>27502<br>27503<br>27506<br>27507<br>27508<br>27509<br>27510<br>27511 | 78.15<br>78.55<br>79.15<br>79.25<br>79.35<br>79.45<br>79.55<br>79.65<br>79.95 | 0QH^*4Z<br>0QHB*4Z<br>0QHC*4Z<br>0QS^*4Z<br>0QSB*4Z<br>0QSC*4Z<br><br>^ = 6, 7, 8, 9<br>* = 0, 3, 4 | 820.2<br>820.3<br>821.+ | S72.10+<br>S72.14+<br>S72.2+<br>S72.3+<br>S72.40+<br>S72.41+<br>S72.42+<br>S72.43+<br>S72.45+<br>S72.46+<br>S72.49+<br>S72.8+<br>S72.9+<br>M97+ | |

| | | | | | | |
|---|---|---|---|---|---|---|
| | | 27513<br>27514 | | | | |
| | Nonunion | 27470<br>27472<br>27170<br>27165 | | | 733.82 | S72.10%@<br>S72.14%@<br>S72.2%X@<br>S72.30%@<br>S72.32%@<br>S72.33%@<br>S72.34%@<br>S72.35%@<br>S72.36%@<br>S72.39%@<br>S72.40%@<br>S72.41%@<br>S72.42%@<br>S72.43%@<br>S72.45%@<br>S72.46%@<br>S72.49%@<br>S72.8X%@<br>S72.9%X@<br><br>%= 0-9<br>@=K, M, N |

| | | | | | | | | |
|---|---|---|---|---|---|---|---|---|
| Tibia & Fibula | Fracture | 27530<br>27535<br>27536<br>27750<br>27752<br>27756<br>27758<br>27759<br>27760<br>27762<br>27766<br>27769<br>27780<br>27781 | 27784<br>27786<br>27788<br>27792<br>27808<br>27810<br>27814<br>27816<br>27818<br>27822<br>27825<br>27826<br>27827<br>27828 | 78.17<br>78.57<br>79.06<br>79.16<br>79.26<br>79.36<br>79.46<br>79.56<br>79.66<br>79.96 | 0QH&*4Z<br>0QH&*5Z<br>0QH&*6Z<br>0QS&*4Z<br>0QS&*5Z<br>0QS&*6Z<br><br>& = G, H, J, K<br>* = 0, 3, 4 | 823.0+<br>823.1+<br>823.2+<br>823.3+<br>823.8+<br>823.9+<br>824.+ | S82.10+<br>S82.12+<br>S82.13+<br>S82.14+<br>S82.16+<br>S82.19+<br>S82.2+<br>S82.30+<br>S82.39+<br>S82.4+<br>S82.5+<br>S82.6+<br>S82.83+<br>S82.84+<br>S82.85+<br>S82.86+<br>S82.87+<br>S82.89+<br>S82.9+ | |
| | Nonunion | 27720<br>27722<br>27724<br>27725<br>27726 | | | | 733.82 | S82.10%@<br>S82.12%@<br>S82.13%@<br>S82.14%@<br>S82.19%@<br>S82.20%@<br>S82.22%@<br>S82.23%@<br>S82.24%@<br>S82.25%@<br>S82.26%@<br>S82.29%@<br>S82.30%@<br>S82.39%@<br>S82.40%@<br><br>%= 0-9<br>@=K, M, N | S82.42%@<br>S82.43%@<br>S82.44%@<br>S82.45%@<br>S82.46%@<br>S82.49%@<br>S82.5%X@<br>S82.6%X@<br>S82.83%@<br>S82.84%@<br>S82.85%@<br>S82.86%@<br>S82.87%@<br>S82.89%@<br>S82.9%X@<br><br>%= 0-9<br>@=K, M, N |

Supplemental Table 2. Beta blocker medications grouped by cardio selective (B1 antagonists) or nonselective (B1 and B2 antagonists).

| **Cardioselective beta blockers** | **Nonselective beta blockers** |
|---|---|
| Atenolol | Propranolol |
| Metoprolol | Nadolol |
| Bisoprolol | Pindolol |
| Nebivolol | Labetalol |
| | Carvedilol |
